# Multi-omics profiling of DNA methylation and gene expression alterations in human cocaine use disorder

**DOI:** 10.1101/2024.04.24.24306302

**Authors:** Eric Zillich, Hanna Belschner, Diana Avetyan, Diego Andrade-Brito, José Jaime Martínez-Magaña, Josef Frank, Naguib Mechawar, Gustavo Turecki, Judit Cabana-Domínguez, Noèlia Fernàndez-Castillo, Bru Cormand, Janitza L. Montalvo-Ortiz, Markus M. Nöthen, Anita C. Hansson, Marcella Rietschel, Rainer Spanagel, Stephanie H. Witt, Lea Zillich

## Abstract

Structural and functional changes of the brain are assumed to contribute to excessive cocaine intake, craving, and relapse in cocaine use disorder (CUD). Epigenetic and transcriptional changes were hypothesized as a molecular basis for CUD-associated brain alterations. Here we performed a multi-omics study of CUD by integrating epigenome-wide methylomic (N=42) and transcriptomic (N=25) data from the same individuals using postmortem brain tissue of Brodmann Area 9 (BA9). Of the N=1,057 differentially expressed genes (p<0.05), one gene, *ZFAND2A*, was significantly upregulated in CUD at transcriptome-wide significance (q<0.05). Differential alternative splicing (AS) analysis revealed N=98 alternatively spliced transcripts enriched in axon and dendrite extension pathways. Strong convergent overlap in CUD-associated expression deregulation was found between our BA9 cohort and independent replication datasets. Epigenomic, transcriptomic, and AS changes in BA9 converged at two genes, *ZBTB4* and *INPP5E.* In pathway analyses, synaptic signaling, neuron morphogenesis, and fatty acid metabolism emerged as the most prominently deregulated biological processes. Drug repositioning analysis revealed glucocorticoid receptor targeting drugs as most potent in reversing the CUD expression profile. Our study highlights the value of multi-omics approaches for an in-depth molecular characterization and provides insights into the relationship between CUD-associated epigenomic and transcriptomic signatures in the human prefrontal cortex.

## Introduction

Cocaine use disorder (CUD) is a globally prevalent substance use disorder (SUD) with around 4.2 million people worldwide being diagnosed with CUD^1^. Individuals suffering from CUD present with compulsive cocaine use patterns, strong cocaine craving, and high rates of relapse even after prolonged time of abstinence^2^. Currently, there is no FDA-approved pharmacotherapy for CUD and treatment is mainly focused on symptom reduction^3^. Neurobiological alterations in the brain are assumed to contribute to the observed clinical symptoms in CUD^4^. This is supported by neuroimaging studies that have shown profound structural and functional alterations in the brain in individuals with CUD^5,6^. In addition to striatal brain regions involved in reward processing^7^, frontal cortical areas that are neuroanatomically connected with limbic structures, are implicated in addiction due to their importance for inhibitory control^5,6,8^.

Dynamic changes in epigenetics and gene expression were hypothesized as a molecular basis of CUD-associated brain changes^9,10^. So far, the majority of studies investigating brain tissue focused on rodent models of cocaine addiction, identifying specific genomic loci to be differentially methylated in brain regions such as the prefrontal cortex (PFC)^11^ and nucleus accumbens (NAc)^12^. Gene expression levels are tightly regulated by epigenetic mechanisms and DNA methylation (DNAm) changes especially in gene promoter regions were shown to alter transcript abundance^13^. In line with this, differential gene expression in rodent models of cocaine addiction was reported in different brain regions where transcription factors of the immediate early gene (IEG) family such as *Egr1*, *Nr4a1*, and *Fos* were found to be differentially expressed^14–17^. At the transcriptome-wide scale, differentially expressed genes were consistently enriched in biological processes related to neurotransmission and ion channel activity, but also metabolic alterations related to lipid metabolism and ATP homeostasis were found^14^.

Few studies have been performed investigating genome-wide DNAm or transcriptomic changes in CUD in human postmortem brain tissue. Two epigenome-wide studies using reduced representation bisulfite sequencing (RRBS) in a cohort of N=25 individuals with CUD and N=25 control individuals identified N=145 and N=173 CUD-associated differentially methylated regions (DMRs) in the nucleus accumbens (NAc)^18^ and in the caudate nucleus (CN)^19^, respectively. Investigating the same brain regions in a different cohort (N=25 CUD cases, N=20 controls), another study characterized transcriptome-wide gene expression changes and reported on the upregulation of synaptic transmembrane transporter genes while immune processes were downregulated^20^. The largest study in the human PFC investigating CUD-associated transcriptomic changes (N=19 CUD, N=17 controls) identified N=883 nominally significant (p<0.05) differentially expressed genes (DEGs) in neuronal nuclei from the Brodmann Area 46 subregion^21^. CUD-associated co-expression networks were enriched for GTPase signaling and neurotransmitter secretion. Regarding epigenomic alterations in the PFC, we were previously able to identify 20 CUD-associated DMRs in Brodmann Area 9, a subregion of the PFC, and further detected that co-methylation networks in CUD were enriched for synaptic signaling processes^22^. Although epigenetics represents an important regulatory mechanism for transcription, the co-regulation of DNAm and gene expression in the same brain samples has not yet been investigated in CUD, limiting the comparability of results between epigenetic and gene expression studies.

In addition to epigenetics and transcription, alterations of alternative splicing might contribute to the neurobiological changes in the CUD brain, as shown in other SUDs. Previous studies using postmortem human brain tissue from individuals with alcohol use disorder (AUD)^23–25^ and opioid use disorder (OUD)^26^ detected differential alternative splicing in transcripts of genes implicated in neuropsychiatric disorders, such as *BIN1*, *FLOT1*, and *ELOVL7* suggesting RNA splicing alterations to be a further molecular mechanism in the neurobiology of SUDs. While a recent study using a cocaine self-administration model in mice showed widespread changes in alternative splicing in multiple brain regions^27^, no systematic evaluation of splicing alterations in human CUD was performed so far.

In the present study, we aimed to characterize the molecular underpinings of CUD in the human prefrontal cortex by applying a multi-omics analysis approach. We investigated differentially expressed genes in postmortem brain tissue from deceased CUD cases compared to well-matched controls and integrated them with the results of our epigenome-wide DNAm analysis^18^ from the same individuals of the BA9 subregion of the human PFC. Further, we characterized differential alternative splicing in BA9. We then performed replication analysis of CUD-associated DEGs in two other independent RNA-seq datasets of human dlPFC. Gene expression data, including alternative splicing results, and DNA methylation data were then integrated and put into a biological context. Finally, we addressed the urgent need for novel therapeutic approaches, by performing a drug repositioning analysis based on the CUD-associated transcriptional profile in BA9.

Collectively, our multi-omics study design represents an integrated analysis of DNAm and gene expression data together with alternative transcript splicing that highlights the role of synaptic and metabolic alterations in CUD and the glucocorticoid receptor as a pharmacological candidate target.

## Results

### Individuals with and without CUD do not differ in sociodemographic characteristics and cell type composition

We first assessed the phenotypic similarities between CUD cases and controls. No significant differences were observed regarding the pH value of the brain, postmortem interval (PMI), RNA integrity number (RIN), and occurrence of comorbid depressive and alcohol use disorders (Supplementary Table S1). We further investigated the variance partition of potential covariates in the RNA-seq dataset (Supplementary Fig. S1a) and found age, PMI, brain pH, and RIN to be associated with gene expression levels, and hence included them as covariates in further analyses. To explore whether major cell type composition could affect analysis results, we performed a cell type deconvolution analysis using CIBERSORT based on human PFC major cell type marker gene signatures (Supplementary Fig. S1b, Supplementary Table S2a). No significant differences in the distribution of major cell types such as astrocytes, oligodentrocytes, microglia, neurons and others were detected between samples from individuals with and without CUD as all 95% high-density intervals from the Bayesian estimation contained 0 (Supplementary Table S2b).

### Transcriptome-wide differential gene expression patterns in CUD are related to synaptic signaling, ion transport, and inflammatory processes

The transcriptome-wide analysis of differential expression in BA9 revealed a total of N=1,057 DEGs associated with CUD (p<0.05). Of these, N=378 were upregulated and N=679 were downregulated (Fig. 1a, Supplementary Table S3). After adjustment for multiple testing, *ZFAND2A* (*Zinc Finger AN1-Type Containing 2A*, log2FC=0.43, p=1.98e-06, q=0.04), remained significantly upregulated in individuals with CUD (5% FDR). We observed a genomic inflation factor of α=1.29 (Supplementary Fig. S1c) and results were stable in a sensitivity analysis without individuals with AUD or MDD (Supplementary Fig. S1d). To evaluate whether BA9 DEGs are significantly enriched within cell-type specific genes of the human PFC, we performed an overlap analysis, using the same set of major brain cell type marker genes as in the cell type composition analysis. Upregulated DEGs were significantly enriched for neuron marker genes exclusively, whereas downregulated DEGs were significantly enriched in markers of non-neuronal cell types such as astrocytes, endothelial cells, and oligodendrocytes (Fig. 1b).

**Fig. 1.**
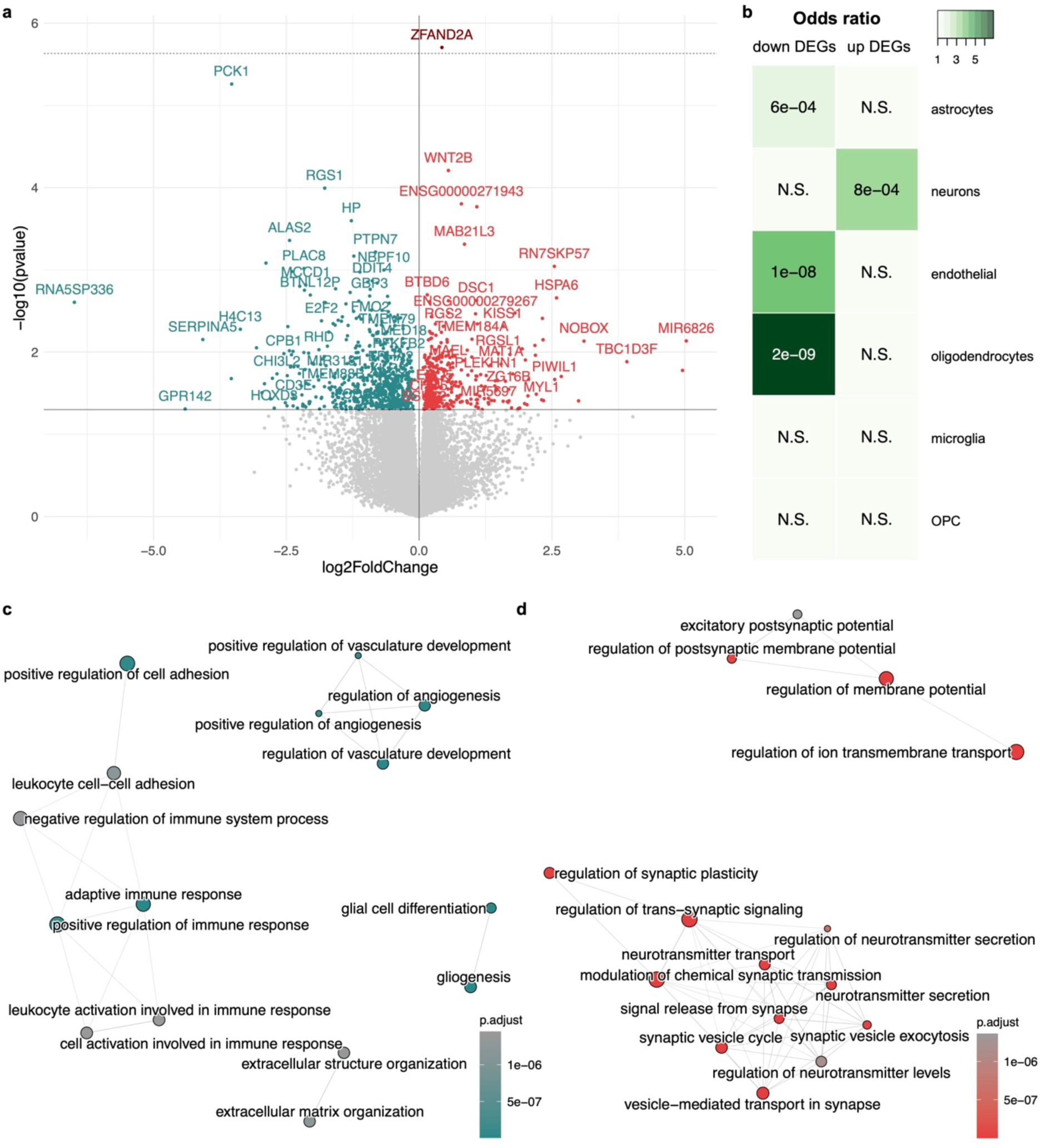
Differential expression analysis in CUD suggests synaptic signaling and immunological alterations in Brodmann Area 9. **a** Volcano plot of the differential expression (DE) analysis showing the N=378 upregulated (red) and N=679 downregulated genes (blue) at nominal significance (p<0.05). Solid black line indicates nominal significance (p<0.05), dashed gray line indicates transcriptome-wide significance (FDR q<0.05). **b** Results of the overlap analysis for upregulated (up) and downregulated (down) DEGs among cell-type specific marker genes. Green color depicts the odds ratio (OR) of overlap, p-values inside the panels indicates significance of overlap based on Fisher-Test. Gene-set enrichment analysis (GSEA) was performed for the DEGs in BA9 ranked by the Wald test statistic from DESeq2. Statistically significant results (q<0.05) from GSEA separated by **c** negative and **d** positive normalized enrichment scores (NES) are shown in an enrichment map visualization. N.S. = not significant, OPC = oligodendrocyte progenitor cell.

Next, we were interested in the biological functions related to the identified CUD-associated DEGs. After adjusting for multiple testing, we detected N=276 statistically significant GO terms for positive GSEA normalized enrichment scores (NES, Supplementary Table S4) and N=782 significant GO terms for negative NES (Supplementary Table S5). Among significantly enriched pathways, the largest positive NES was detected for “vesicle-mediated transport in synapse” (NES=2.63, q=5.31e-15), whereas “superoxide metabolic process” (NES=-2.43, q=2.08e-06) was the top finding with negative NES. To identify functional modules of pathways consisting of multiple GO terms related to similar biological functions, we created an enrichment map (emap) visualization based on the significant findings from GSEA. For GO terms with negative NES, we detected one large cluster related to inflammatory and immune signaling and several smaller clusters consisting of pathways involved in angiogenesis, extracellular matrix (ECM) organization, and gliogenesis (Fig. 1c). Two major clusters emerged for pathways with positive NES. The first was related to neurotransmission and synaptic signaling whereas the second cluster consisted of GO terms involved in transmembrane transporter activity (Fig. 1d).

### Network analysis highlights fatty-acid metabolism and morphogenesis processes in CUD

We next performed weighted gene co-expression network analysis (WGCNA) to investigate gene co-expression patterns in CUD and detected a total of N=27 co-expression modules (Supplementary Fig. S2a+2b). Co-expression module yellow was significantly correlated with CUD (r=-0.47, p=0.02) while no significant association with other known covariates was observed (Supplementary Fig. S2a). Module yellow consisted of N=2,517 genes and module membership was highly correlated with gene significance for CUD (r=0.61, p<1e-200, Supplementary Fig. S2c, Table S6). GO enrichment analysis for module yellow genes revealed N=519 statistically significant GO terms after multiple testing correction (Supplementary Table S7). Strongest associations were detected for “small molecule catabolic process” (q=2.22e-11), and more specifically, “carboxylic acid catabolic process” (q=1.59e-09, Supplementary Fig. S2d). After clustering the significant terms, a prominent GO term cluster related to fatty acid metabolism was detected, while another cluster was related to organ developmental and morphogenesis processes. To further characterize WGCNA expression module yellow, we generated a protein-protein interaction (PPI) network based on module hub genes and identified APOE (N=9 edges), ERBB2 (N=8 edges), ALDH7A1 (N=7 edges), PPARA (N=7 edges), and TLR4 (N=7 edges) as the most strongly connected nodes in the PPI network (Supplementary Fig. S3, Supplementary Table S6).

### Genes with alternative splicing events in CUD are involved in cell junction formation and the morphogenesis of axons and dendrites

To investigate alternative splicing in CUD and its potential relevance for contributing to altered neurobiological functions in the brain, we performed a differential alternative splicing analysis using LeafCutter^28^. After multiple testing correction, we identified a total of N=108 differentially spliced intron clusters in BA9 (FDR<0.05, Fig. 2a, Supplementary Tables S8-S10). These clusters were distributed among N=98 genes that we further denote as alternatively spliced (AS) genes. One of the top findings in our AS analysis of CUD was *BIN1* (*Bridging Integrator 1*, q=7.8e-04, Fig. 2b) previously identified as a conserved AS genes in other substance use disorders. We next investigated the biological pathways enriched for alternative splicing events based on our list of AS genes. Statistically significant enrichment after multiple testing correction was detected for N=15 GO terms (Supplementary Table S10). Strongest enrichment was found for GO terms “cell junction assembly” and “neuron projection extension” (both q=3.62e-03). In the emap visualization of enriched GO terms with a more lenient threshold of 25% FDR (q<0.25), we detected a well-connected cluster containing GO terms related to cellular growth and cell-cell junction development, while also more brain-specific processes such as myelination and the extension of axons and dendrites were found (Fig. 2c). While differential alternative splicing itself contributes to altered biological functions by inducing different abundances of transcript isoforms, this effect might be potentiated by differential gene expression. We thus investigated the overlap of AS and DEGs in CUD and identified 8 genes that were differentially spliced and differentially expressed in BA9: *ITPKB, CPLX1, HLA-F, INPP5E, GALNT8, IGFBP6, ZBTB4,* and *BCAT2* (Fig. 2d).

**Fig. 2.**
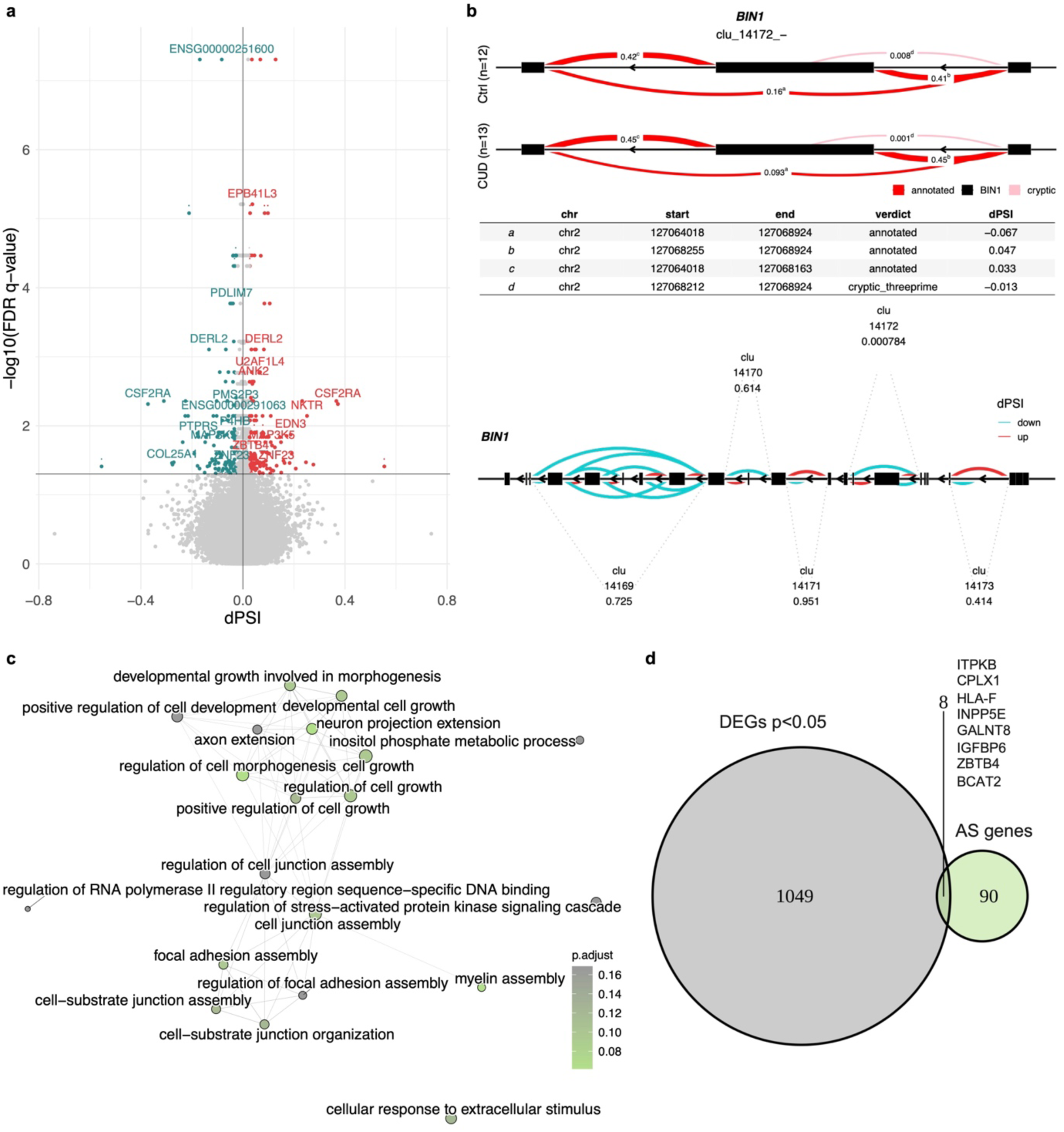
Differential alternatively spliced genes in CUD are related to neuron morphogenetic processes. **a** Volcano plot of the differential alternative splicing (AS) results in Brodmann Area 9 (BA9). Statistically significant intron clusters (N=108) identified by LeafCutter (q<0.05) were annotated by gene name while dots represent individual introns of an intron cluster. Introns highlighted in red (dPSI>0) are more abundant in CUD while introns highlighted in blue (dPSI<0) are less abundant in CUD. **b** Results of the differential AS analysis at the cluster and gene level for one of the top findings, an intron cluster (clu_14172_-) in the Bridging Integrator 1 (*BIN1*) gene. Upper panel: visualization of *BIN1* exons and introns with percent spliced in (PSI) measures related to the significant cluster clu_14172_-. The table indicates delta percent spliced in (dPSI) values from the CUD vs. Ctrl comparison. Lower panel: gene-level summary of all intron clusters detected in *BIN1*. FDR q-values are shown below cluster names. **c** GO enrichment analysis for the N=98 AS genes harboring differentially statistically significant (q<0.05) intron clusters in CUD. **d** Overlap of findings from differential expression (DE) analysis (N=1057 DEGs at p<0.05) and differential AS analysis.

### Replication analysis in independent cohorts reveals *FKBP4* and *HSPA6* as conserved DEGs in CUD

To evaluate the potential replication of CUD-associated DEGs in other RNA-seq datasets of human PFC, we performed an overlap analysis of nominally significant DEGs (p<0.05) across studies. CUD-associated differential expression testing was performed in two independent replication datasets, the first originating from BA9 (BA9 replication, bulk RNA-seq) and the second from BA46 (BA46 replication, neuron-specific RNA-seq). Two genes, *HSPA6* and *FKBP4*, were shared upregulated DEGs at nominal significance and showed comparable effect sizes (log2FC) in CUD across all three PFC datasets (Fig. 3a+c). As *HSPA6* is a spliceosome-associated gene with conserved differential expression across datasets, we performed a look-up of genes related to the KEGG Spliceosome pathway (hsa03040) in DE results from our discovery cohort (Fig. 3b). Here, we aimed to address the hypothesis of spliceosomal differential gene expression as a potential mechanism for splicing alterations in CUD^29^. *HSPA6* was the spliceosome-associated gene showing strongest CUD-associated expression changes in BA9 (log2FC=2.59, p=0.002). We detected six additional spliceosome-associated genes that were among nominally significant DEGs: *HSPA1A* (log2FC=0.71, p=0.034), *CRNKL1* (log2FC=-0.19, p=0.011), *LSM6* (log2FC=-0.24, p=0.014), *SRSF4* (log2FC=-0.14, p=0.022), *SNRPG* (log2FC=-0.20, p=0.037), and *TRA2A* (log2FC=-0.14, p=0.036).

**Fig. 3.**
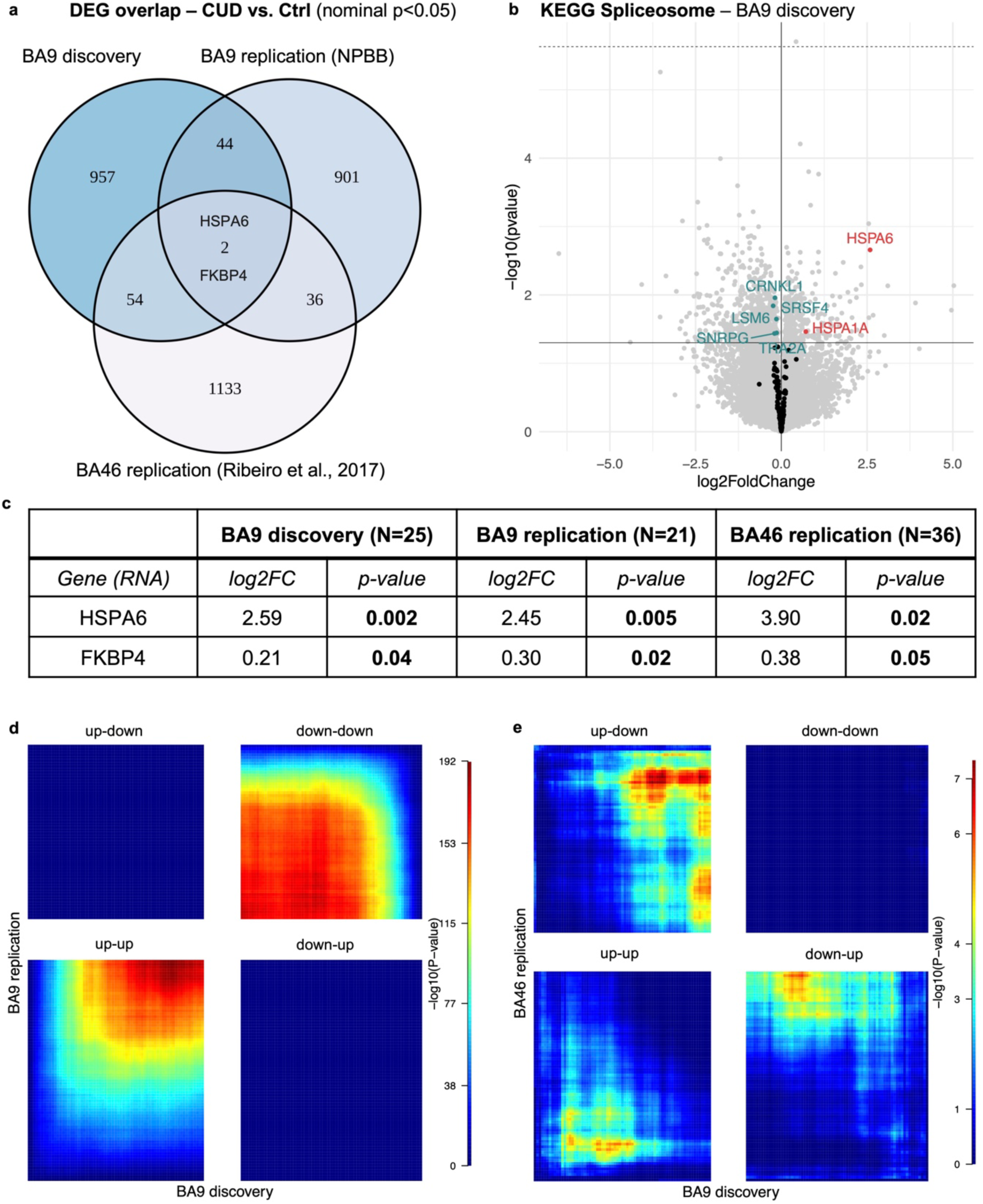
Replication analysis of CUD associated transcriptomic alterations in independent datasets. **a** Overlap of nominally significant (p<0.05) differentially expressed genes across datasets reveals two shared DEGs, HSPA6 and FKBP4. The replication datasets were based on N=21 BA9 samples from the National PTSD Brain Bank (NPBB) and neuronal-specific transcriptomic data of N=36 BA46 samples available under GEO accession number GSE99349 (BA46 replication). **b** HSPA6 is the strongest spliceosome-associated DEG in BA9. **c** Results of the look-up analysis for shared DEGs HSPA6 and FKBP4 - log2-fold change and p-value: association p-value from the DESeq2-based differential expression results. Significant associations are highlighted in bold. Rank-rank hypergeometric overlap (RRHO) visualization for **d** the BA9 replication dataset and **e** the neuronal-specific BA46 dataset indicating convergent and divergent expression patterns across studies using full differential expression statistics as the input datasets. Color scale represents -log10(p) of the hypergeometric testing procedure in RRHO. Convergent expression across datasets: up-up and down-down, divergent: up-down and down-up.

Using rank-rank hypergeometric overlap (RRHO) visualization for a more unbiased evaluation of convergent and divergent gene expression patterns across studies, we found strong convergent overlap between the BA9 discovery and BA9 replication datasets indicating similar patterns of CUD-associated expression deregulation (Fig. 3d). In the comparison with neuron-specific expression data from BA46, we found prominent divergent gene expression patterns between datasets, while convergent expression patterns were enriched in the shared upregulated genes across studies (Fig. 3e).

### Drug repositioning analysis highlights glucocorticoid receptor targeting drugs to reverse the CUD gene expression profile

To evaluate the potential use of small molecule drugs to revert the gene expression pattern of CUD, we performed drug repositioning analysis based on the L1000 assay as implemented in CMap (Supplementary Fig. S4a), using the top 150 up- and downregulated genes as input (Supplementary Table S11). Among the results with negative normalized connectivity score (NCS), i.e. perturbagens that revert the DE profile in CUD, the most significant finding for small molecule drugs after multiple testing correction was the glucocorticoid receptor agonist medrysone (NCS=-1.78, q=2.2e-16, Supplementary Fig. S4b). Glucocorticoid receptor agonists were the only FDR-significant perturbagen class overrepresented among CMap GSEA results (Supplementary Fig. S4c). When we further investigated connectivity scores for all glucocorticoid receptor targeting drugs including agonists and antagonists in CMap, we found exclusively significant negative connectivity scores (Supplementary Fig. S4d) suggesting glucocorticoid receptor targeting molecules as potential pharmacological drugs to revert the CUD expression changes in BA9. In line with this finding, the biological pathway “response to glucocorticoid” (NES=-1.54, q=0.019, Supplementary Table S5) was among the FDR-significant pathways with negative NES in the GSEA analysis of DEGs from BA9.

### Findings of the integrated analysis of DNA methylation and gene expression data converge at the gene and pathway levels

As DNA methylation data was available and previously analyzed for the same cohort in BA9, we next aimed to integrate findings from the epigenome-wide and transcriptome-wide studies on the gene-level, applied multi-omics factor analysis, and performed an integrative functional GO-term enrichment analysis across all –omics layers.

### Gene-level integration of epigenomic, transcriptomic and splicing alterations highlights ZBTB4 and INPP5E in CUD

Of the overlapping genes between the differential methylation, expression, and alternative splicing analyses, two genes were consistently altered across all the investigated molecular views in BA9: *ZBTB4 (Zinc Finger And BTB Domain Containing 4)* and *INPP5E (Inositol polyphosphate-5-phosphatase E)* (Fig. 4a, Supplementary Table S12). Both genes were characterized by a hypomethylated CpG site and increased transcript levels in CUD (Fig. 4b). For *ZBTB4*, the strongest association for a CpG site was found for cg03443505 (chr17:7387573, β=-0.84, p=1.01e-05). *ZBTB4* was upregulated with a log2FC of 0.08 (p=0.015) and it contained the differentially spliced intron cluster *chr17:clu_10246_-* (q=0.028). The strongest association for CpG differential methylation in the *INPP5E* gene was found for cg18558462 (chr9:139334381, β=-0.93, p=8.55e-03). It was differentially expressed with log2FC of 0.17 (p=0.025) and intron cluster *chr9:clu_25078_-* was differentially alternatively spliced (q=0.015). In the replication datasets, we detected conserved transcript upregulation of *ZBTB4* (BA9 replication, log2FC=0.12, pval=0.21; BA46, log2FC=0.12, pval=0.09) and *INPP5E* (BA9 replication, log2FC=0.11, pval=0.47; BA46, log2FC=0.08, pval=0.49), however not statistically significant. To deeper characterize the *ZBTB4* and *INPP5E* gene loci in BA9 and specifically in the context of CUD, we performed an integrative gene locus visualization approach by combining GWAS, EWAS, alternative splicing, and RNA-seq results for CUD with ENCODE ChIP-seq reference data from human dorsolateral prefrontal cortex. ChIP-seq data confirmed the presence of activating chromatin marks at promoter (H3K4me4, H3K27ac) and gene body regions (H3K36me3) at *ZBTB4* and *INPP5E* gene loci in the human dlPFC. In addition, for *ZBTB4*, multiple nominally significant associations for SNPs and CpG sites were detected that were most prominent within intronic and intergenic regions, while no SNP but CUD-associated CpG sites were identified in the *INPP5E* gene locus (Fig. 4c). In line with this, when we quantified the association of genetic variants with CUD at the gene level using a gene-based association analysis in MAGMA, we detected stronger association for *ZBTB4* with CUD (Z=1.74; p=0.04) compared to *INPP5E* (Z=-1.14; p=0.87).

**Fig. 4.**
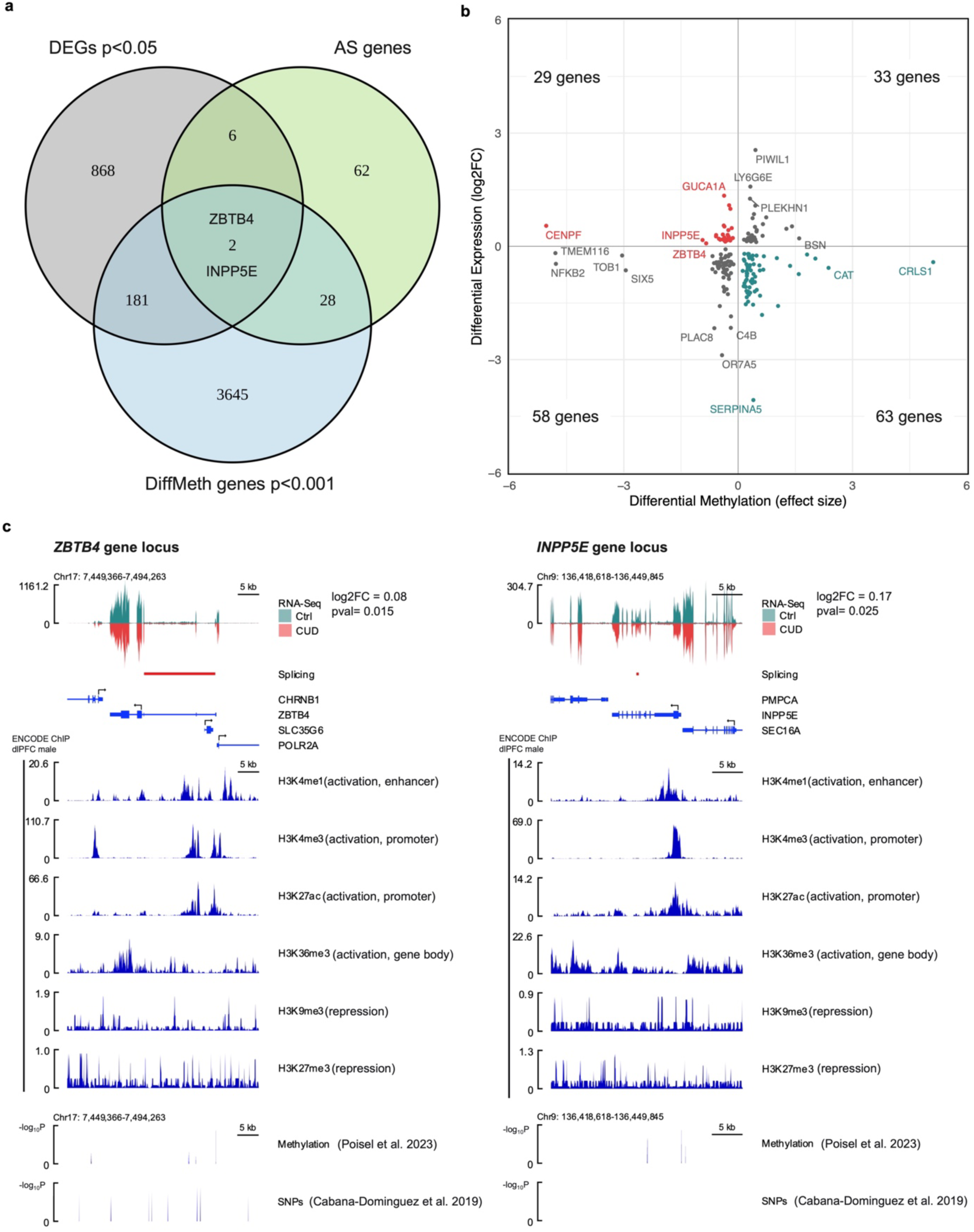
Convergence of DNA methylation, alternative splicing, and gene expression alterations in CUD at the *ZBTB4* and *INPP5E* gene loci. **a** Overlap of differential expression (DE), differential DNA methylation (DiffMeth), and differential alternative splicing analyses suggest two genes, *ZBTB4* and *INPP5E*, where alterations are consistently associated with CUD. **b** Relationship between DE and DiffMeth genes in Brodmann Area 9 based on log2FC (y-axis) from DE analysis and effect size β from linear regression in the EWAS of CUD (x-axis). For both genes, *ZBTB4* and *INPP5E* (highlighted in red), hypomethylation of the strongest significant CUD-associated CpG site and increased transcript levels are observed. **c** Integrated visualization of functional genomic datasets for *ZBTB4* and *INPP5E* gene loci. CUD-associated genomic variants (SNPs p<0.05 from^30^), CUD-associated CpG sites (p<0.05 from^22^), RNA-seq data and intron clusters (q<0.05) from the present study were visualized together with ENCODE ChIP-seq data for different chromatin marks in human dorsolateral prefrontal cortex.

### Multi-Omics Factor Analysis confirms cell junction, synaptic signaling, and neurogenesis as important biological processes in CUD

The integration of DNA methylation and gene expression data as described above is based on results of the EWAS, DE, and AS analyses which is one possible way of integrating multiple omics datasets. In addition, multi-omics analysis tools such as MOFA enable an integrated analysis of omics datasets in a single statistical framework. Using MOFA on our DNA methylation and gene expression data from BA9, we identified one factor representation of the multi-omics dataset (factor 9) that was significantly correlated with CUD (r=-0.48, p=0.02) and age (r=0.47, p=0.02, Supplementary Fig. S5a+S5b). Factor 9 displayed significantly smaller factor values in CUD cases compared to individuals without CUD in a Wilcoxon test (p=0.02, Supplementary Fig. S5c). When we extracted the CpG sites with the strongest weights on factor 9, cg23859635 annotated to *MTA3* was the CpG site with the strongest positive weight on factor 9 (w=0.31), while cg24621354 in the gene *TES* displayed the strongest negative weight (w=- 0.33, Supplementary Fig. S5d, Supplementary Table S13). In the gene expression dataset, the small GTPase *RAB6A* had the strongest positive weight (w=0.07), while *HIVEP2* had the strongest negative weight (w=-0.05) on factor 9 (Supplementary Fig. S5e, Table S13). Results of a GSEA on negative expression weights on factor 9 revealed FDR-significant (q<0.05) enrichment for synaptic signaling, cell junction organization, and neurogenesis pathways, confirming the results from the previous analyses. In contrast, GSEA on positive expression weights revealed enrichment for cellular respiration and small molecule metabolic processes (Supplementary Fig. S5f, Supplementary Table S14). When we used missMethyl to investigate the biological pathways for DNA methylation features with strong weights on factor 9, we detected enrichment for similar biological pathways as in the analysis of expression features. While none of the enrichment results remained FDR-significant after multiple testing correction, strongest enrichment for CpG sites with negative weights on factor 9 was detected for intracellular calcium concentration regulation and synaptic vesicle related processes (Supplementary Fig. S5g, Supplementary Table S15). The pathways showing the strongest enrichment for the positive weight CpG sites were related to monocarboxylic acid and specifically, lactate transmembrane transporter activity, and ER stress pathways (Supplementary Fig. S5h, Supplementary Table S15).

### Integrative functional analysis reveals functional modules related to neurotransmission, cell differentiation, cell junction organization, and fatty acid metabolism

In an integrative functional analysis approach, we used all available information from our study on DNA methylation, gene expression, and alternative splicing alterations in CUD to identify potential convergence of association results at the pathway level in BA9. We thus performed a GO enrichment analysis based on 10 curated lists with CUD-associated genes derived from EWAS, DE analysis, alternative splicing analysis, WGCNA modules based on DNA methylation and expression data, and MOFA (Supplementary Table S16). In the enrichment map for GO terms, we identified several functional modules where the same biological pathway was detected for multiple gene lists at FDR-adjusted statistical significance (q<0.05) indicating convergence of the results from different analysis approaches (Supplementary Fig. S6). The two largest functional modules (FM) contained pathways involved in neurotransmission and synaptic signaling (FM1), while FM2 was enriched for neuron and glial cell differentiation, growth, and morphogenesis processes. Two further prominent modules were related to synapse and cell junction organization (FM3) and fatty acid metabolism (FM4).

## Discussion

By applying a multi-omics data integration approach on DNA methylation and gene expression data from postmortem human brain tissue we aimed for a deeper understanding of the neurobiology of CUD in the human prefrontal cortex. At the gene level, our differential expression analysis suggests two candidates, *FKBP4* and *HSPA6*, which were replicated as nominally significancant findings in two independent cohorts. In addition, our multi-omics analyses highlight *ZBTB4* and *INPP5E*, that were consistently altered across omics analyses in BA9 and displayed consistent upregulation patterns in independent replication datasets. At the pathway level, we found converging evidence for CUD-associated DNAm and transcriptional alterations that were related to neurotransmission, fatty-acid metabolism, and changes in neuronal morphology.

Analysis of the transcriptome in BA9 revealed *ZFAND2A* as the DEG showing the strongest association with CUD. *ZFAND2A* is a canonical heat shock gene in humans encoding a zinc-finger containing protein that is involved in the regulation of proteasomal protein degradation^31,32^. It was further identified as a DEG in a study on transcriptomic signatures of Alzheimeŕs disease (AD)^33^. Another AD-related finding emerged in co-expression network analysis. APOE showed the strongest connectivity in the PPI network for module yellow genes and has been intensively characterized due to its association with age of onset in AD^34^. While SUDs and neurodegenerative disorders such as AD depict different neuropsychiatric disorders based on the current understanding of disease mechanisms, CUD and AD share brain atrophic changes as a clinical symptom^35^ and our results suggest that there might be shared molecular mechanisms involved.

Previous studies have identified differential alternative splicing in alcohol use disorder^25^ and opioid use disorder (OUD)^26^ in the human brain, however, RNA splicing alterations have not been characterized in human CUD so far. In the differential alternative splicing analysis, we found N=98 statistically significant genes containing AS intron clusters. Interestingly, among our top findings, we found *Bridging Integrator 1* (*BIN1*) for which differential alternative splicing in the brain has been described in OUD. *BIN1* was the only differential AS gene in OUD that was conserved across all investigated brain regions; dlPFC, NAc, and midbrain^26^. Further, AS events in *Bin1* were identified in the mouse brain in a study on splicing alterations associated with cocaine self-administration^27^. As dendrite and axon morphogenesis processes were among the enrichment results for AS genes in BA9, we hypothesize that AS is directly related to neuroplastic changes in the CUD brain. Mechanistically, AS processes change the abundance of transcript isoforms with different biological functions that might contribute to the neuroadaptations in CUD. We explored the mechanism of spliceosomal gene expression alterations as a potential contributor to differential AS events in CUD. Exposure to cocaine was previously hypothesized to alter spliceosomal gene expression^29^ and our results suggest spliceosomal genes such as *HSPA6* and *HSP1A1* as DEGs in BA9. As spliceosomal gene alterations were also detected in the replication analysis with *HSPA6* as a shared upregulated DE gene across studies, AS events might be an important mechanism in CUD contributing to neurobiological changes in the PFC.

In the last step of the RNA-sequencing analysis, we aimed to address the urgent need for novel pharmacotherapeutic approaches for the treatment of CUD by performing a drug repositioning analysis. We detected glucocorticoid receptor-targeting drugs having consistently negative connections with the CUD expression profile in BA9. In addition, *FKBP4*, an important regulator of glucocorticoid receptor signaling was identified as a conserved upregulated DEG in CUD based on three independent dlPFC datasets. FKBP4 has a key role in the nuclear translocation of the glucocorticoid receptor, as it replaces FKBP5 upon cortisol binding to the receptor complex leading to its nuclear translocation^36^.

Pharmacological targeting of glucocorticoid receptor signaling was tested in rodent models of cocaine addiction ^37–39^. Reduced behavioral response to cocaine was observed when glucocorticoid receptor antagonists such as mifepristone were applied^37^. In contrast, corticosterone was shown to promote cocaine intake in rats^38,39^. In the drug repositioning analysis, results for glucocorticoid receptor agonists were more prominent compared to antagonists which appears to be in conflict with previous literature. However, glucocorticoid receptor antagonists such as mifepristone also displayed significant negative connectivity scores with the BA9 expression signature supporting previous findings. Further, synthetic glucocorticoid receptor agonists such as dexamethasone were shown to impair cocaine self-administration in rats^38^ indicating a more complex relationship between the endogenous glucocorticoid system and exogenously applied glucocorticoid receptor targeting drugs. We thus suggest that glucocorticoid receptor targeting drugs should be further investigated for their potential use as a pharmacotherapy in CUD.

Using multi-omics data integration, we identified two genes, *ZBTB4* and *INPP5E,* for which CUD-associated alterations were consistently detected across DNAm, gene expression, and alternative splicing analyses. Both genes contained a hypomethylated CpG site, stronger transcript expression was found in individuals with CUD, and significant differentially spliced intron clusters were identified. Despite being strongly expressed in the brain and most prominently in neurons^40^, the role of *ZBTB4* in neuropsychiatric disorders remains poorly understood. However, due to the DNA binding capacity and its role as a transcriptional repressor, *ZBTB4* deregulation in CUD could lead to downstream expression changes of its target genes. Further, protein-protein interaction data suggests interaction of ZBTB4 with the transcription factor PRDM5 as well as with the AP2M1 and AP2A1 subunits of the adapter protein 2 (AP-2) complex that is involved in endocytosis of neurotransmitter receptors in neurons^41,42^. The second finding at the gene level, *INPP5E*, encodes a phosphatidylinositol-phosphatase specific to cilia and *INPP5E* mutations were found in Joubert syndrome which is characterized by cerebellar and cerebral malformation^43^. A possible link to CUD provide neuronal primary cilia, known as key signaling hubs on somata enriched for G-protein-coupled receptors (GPCRs)^44^. As INPP5E is required for proper trafficking of GPCRs along ciliary microtubules^45^, deregulation of *INPP5E* might lead to aberrant ciliary signaling that has recently gained attention in the addiction field: cell type-specific ablation of neuronal primary cilia in mice was shown to affect body weight as well as locomotor response to psychostimulants such as cocaine^46^ and amphetamine^47^. In humans, further studies on INPP5E are required to evaluate its role in SUDs.

Evaluating the convergence of results at the pathway level revealed widespread molecular alterations in synaptic signaling represented by functional module FM1 in the GO enrichment analysis. Our findings are well in line with previous literature that reported on cocaine-associated DNAm and expression changes in genes involved in neurotransmission^14–16,18,48^. The observed overrepresentation of neuronal marker genes in the upregulated DEGs together with the non-neuronal marker gene enrichment in the downregulated DEGs further suggests a particular importance of CUD-associated expression changes in altering neurotransmission. In a study on CUD-associated gene expression changes in neuronal nuclei of the human dlPFC^21^, the authors found a WGCNA co-expression module that was significantly associated with CUD and was enriched for GTPase signaling and neurotransmitter transport that well matches our results in BA9. Neuronal function thus appears to be strongly influenced by altered epigenetic and transcriptional programs in the CUD brain.

Functional modules FM2 and FM3 were related to pathways involved in neuron, synapse, and axon morphogenetic processes. This is supported by literature from animal models of CUD, where alterations in dendritic branches and spine density were observed in the PFC of cocaine self-administering rats^49^. Even a single cocaine exposure was sufficient to reduce dendritic spine density in neurons^50^. In summary, brain morphological changes depict an interesting link between molecular and behavioral aspects of addiction as neuroplastic changes are the basis of neurocircuit alterations in the SUD brain that are related to compulsive drug-seeking and relapse^51^.

Another converging finding were metabolic changes related to fatty acid metabolism (FM4). This finding was especially prominent in the CUD-associated WGCNA module yellow where we found a functional module of pathways related to fatty acid metabolism. Further, results from MOFA suggested gene expression changes related to the electron transport chain as another key metabolic pathway alteration. This is supported by findings from animal models of cocaine addiction where a downregulation of glycolysis and oxidative phosphorylation were observed in the brain^52^ while fatty acid metabolism genes were upregulated^53^. It has to be noted that metabolic changes in CUD are most likely not brain-spercific but also appear on a systemic level as individuals with CUD were found to have reduced body fat in comparison to a healthy control group^54^. To follow up on this finding, future studies should evaluate if interfering with fatty acid metabolism could depict a therapeutic strategy in CUD as a ketogenic diet has been shown to alter the behavioral response to cocaine in rats^55^.

There are some limitations that apply to our multi-omics study of CUD. First, depicting an inherent limitation of analyses in human postmortem brain tissue, our cross-sectional analysis design can only reflect the endpoint of CUD limiting the identification of dynamic changes in DNAm and gene expression during the disease course. Second, considering the sample size and the few DEGs at transcriptome-wide significance, it remains unclear whether the findings are generalizable to the general population highlighting the need for studies in larger and more diverse cohorts. Third, while the homogeneity of our sample consisting of only males from EA ancestry is a strength in statistical analysis, sex-specific and ancestry-related molecular signatures of CUD remain an open question. At least in the analysis of the more diverse replication cohorts we were able to show comparable CUD-associatied gene expression patterns when compared to our discovery cohort.

In summary, our study identifies novel associations with CUD at the gene level, confirms these on the multi-omics level, and suggests differential alterative splicing as an important molecular hallmark of CUD in the human prefrontal cortex. At the same time, our study supports previous findings of synaptic signaling alterations that have been robustly detected when investigating the neurobiological effects of cocaine. We highlight drugs targeting glucocorticoid receptor signaling to be further tested as a treatment for CUD.

## Author contributions

Conceptualization, E.Z., L.Z., M.R., R.S., and S.H.W.; Methodology, E.Z., H.B., J.F., and L.Z.; Resources, G.T., N.M., A.C.H., J.C-D., N.F-C., B.C., and J.M.O.; Data curation, E.Z., D.A., H.B., D.A-B., J.J.M-M., and L.Z. ; Data Analysis, E.Z., H.B., D.A., D.A-B., L.Z., and J.M.M-M., Investigation, E.Z., H.B., A.C.H., M.R., R.S., S.H.W, and L.Z.; Writing – Original Draft, E.Z., H.B.; Writing – Reviewing & Editing, E.Z., H.B., D.A., D.A-B, J.M-M., J.F., N.M., G.T., J.C-D., N.F-C., B.C., J.L.M-M., A.C.H., M.R., R.S., S.H.W., and L.Z.; Supervision, J.F., M.R., and R.S., J.M.O., L.Z., and S.H.W; Project Administration, R.S., A.C.H., M.R. and S.H.W., Funding Acquisition, R.S., A.C.H., M.R., and S.H.W.

## Competing interests

The authors declare that there are no competing interests.

## Data availability statement

Raw methylation data is deposited in the European Genome Phenome Archive (EGA) under accession number EGAS00001006828 (https://ega-archive.org/studies/EGAS00001006826). RNA-sequencing data has been uploaded to the EGA will be listed with an accession number by the time of publication.

## Code availability statement

All original code used for data analysis and figure preparation will be available in a GitHub repository (https://github.com/lzillich/BA9_multi_omics) by the time of publication.

## Ethics statement

Postmortem human brain tissue was obtained from the Douglas Bell Canada Brain Bank where tissue sampling is performed based on their established ethical guidelines. Our multi-omics study was approved by the Ethics Committee II of the University of Heidelberg, Medical Faculty Mannheim, Germany, under the register number 2021-681.

## Funding

Funding supporting this study was provided by the German Federal Ministry of Education and Research (BMBF) within the e:Med research program SysMedSUDs: “A systems-medicine approach toward distinct and shared resilience and pathological mechanisms of substance use disorders” (01ZX01909 to R.S., M.R., A.C.H., and S.H.W.). In addition, by the Deutsche Forschungsgemeinschaft (DFG) through the collaborative research centre TRR265: “Losing and Regaining Control over Drug Intake”^56^ (Project ID 402170461 to S.H.W., R.S., A.C.H. and M.R.), the Hetzler Foundation for Addiction Research (to A.C.H.), the ERA-NET program: Psi-Alc (01EZ1908), Spanish ‘Ministerio de Ciencia, Innovación y Universidades’ (PID2021-1277760B-I100, to B.C.), ‘Generalitat de Catalunya/AGAUR’ (2021-SGR-01093, to B.C.), ICREA Academia 2021, ‘Fundació La Marató de TV3′ (202218-31, to B.C.) and from ‘Ministerio de Sanidad, Servicios Sociales e Igualdad/Plan Nacional Sobre Drogas’ (PNSD-2020I042, to N.F.-C.). The project has been carried out using the Mannheim (CIMH) infrastructure of the German Center for Mental Health (DZPG).

## Methods

### Postmortem human brain tissue

The sample of human postmortem brain tissue of BA9 was obtained from the Douglas Bell Canada Brain Bank (DBCBB). Inclusion criteria were age > 18 and a diagnosis of cocaine dependence based on DSM-IV. Throughout this study, we will nevertheless use the more recent terminology from DSM-5 i.e., cocaine use disorder. Individuals were excluded from the study if they were diagnosed with severe neurodevelopmental or psychiatric disorders other than depressive disorders or had received additional diagnoses of substance use disorders other than alcohol use disorder. All included subjects were male and of European American descent. Demographic information for the cohort of N=42 BA9 tissue donors is described in^22^ and for the subset of N=25 individuals with RNA-seq data in Table S1.

### DNA methylation data generation

DNA extraction was performed as described in^22^. In brief, DNA was extracted from the full set of N=42 BA9 samples using the DNeasy Blood and Tissue Kit (Qiagen, Hilden, Germany). The epigenome-wide DNAm profile was determined using the Illumina MethylationEPIC BeadChip v1 (850k) (Illumina, San Diego, CA, USA). During sample processing and analysis of DNAm levels, randomization was applied based on CUD status and known comorbidities such as AUD and depressive disorders.

### Generation of gene expression data

Using the miRNAeasy Mini extraction Kit (Qiagen, Hilden, Germany), total RNA was extracted from the N=42 BA9 samples using ∼5mg of frozen tissue from each individual. The RNA integrity number (RIN) was measured using a TapeStation 4200 (Agilent, Santa Clara, CA, USA) resulting in a total of N=25 samples remaining for RNA sequencing (RIN > 5.5). Following ribosomal RNA (rRNA) depletion, libraries were prepared using the NEBNext Ultra II Directional RNA Library Prep Kit (New England Biolabs, Ipswich, MA, USA) followed by sequencing with an average of 60 million read pairs (2x100bp) per sample. RNA sequencing was performed using an Illumina NovaSeq 6000 device.

### Statistical analyses

All statistical analyses in the R programming environment were performed using R version 4.2.1. If not otherwise stated, adjustment for multiple testing was performed using the Benjamini-Hochberg (FDR) procedure^57^. An analysis workflow for the multi-omics study of DNA methylation and gene expression in CUD is shown in Supplementary Fig. S7.

### DNA methylation analysis

Methylation data was analyzed as part of the Poisel, et al. ^22^ study where a detailed description of the analysis pipeline can be found in the methods section. In brief, DNA methylation levels were preprocessed using an in-house quality control (QC) pipeline based on CPACOR^58^. The neuronal cell fraction was estimated based on the Houseman algorithm^59^ using a dlPFC reference dataset^60^. Quantile-normalized beta values were derived from raw-intensities, followed by logit-transformation to M values of methylation. An epigenome-wide association study (EWAS) was performed using a linear regression model while adjusting for covariates that have a known effect on DNA methylation such as age, postmortem interval (PMI), pH of the brain tissue, neuronal cell fraction, comorbid depressive and/or alcohol use disorder, and technical factors. Downstream analyses based on the results of the EWAS included the identification of differentially methylated regions (DMRs), a gene ontology enrichment analysis using CUD-associated CpG sites (p_assoc_<0.001), and a network analysis in WGCNA to evaluate CUD-associated co-methylation modules.

### Gene expression analysis

Sequencing quality metrics were inspected using FastQC v.0.12.1 confirming all 25 fastq files to be used in further analysis. Reads were mapped to the GRCh38 genome primary assembly using STAR v.2.7.10b^61^. Quantification of features was performed using the featureCounts implementation in the R package Rsubread v.2.12.3^62^ with the genome annotation gtf-file v.43 from GENCODE (https://www.gencodegenes.org). The raw count matrix was imported in DESeq2 v.1.38.3^63^ and differential expression (DE) testing was performed while adjusting for the covariates age, PMI, brain pH and RIN in the DESeq2 experimental design formula. The distribution of resulting p-values was assessed in a quantile-quantile plot to evaluate potential genomic inflation (Fig. S1A). Fold-change cut-offs for DEGs were an absolute log2 fold change of larger than 0.07, corresponding to a 5% change in transcript abundance. Statistical significance cut-offs were p<0.05 for nominal significance and q<0.05 for a 5% FDR-adjusted significant association with CUD. All covariates included in the DESeq2 model are known to influence the gene expression profile and were confirmed in a variance partition analysis in our dataset using the R package variancePartition v.1.28.7 (Supplementary Fig. S1a). As comorbid MDD and AUD explained only minimal variance in the expression data and only 25 of the 42 samples were available in the expression analysis, MDD and AUD were not included as covariates in the statistical model. A sensitivity analysis was performed including MDD and AUD as covariates (Supplementary Fig. S1b) confirming a strong correlation between the log2 fold-changes of the nominally significant results.

### Cell-type deconvolution analysis

Based on reference signatures of gene expression derived from single-cell studies, the distribution of cell types in bulk expression data can be inferred using cell-type deconvolution algorithms such as CIBERSORT^64^. We used a curated set of cell type-specific marker genes of the human prefrontal cortex based on a study from Yu and He ^65^ where a gene was required to have a 10-fold stronger expression in a specific cell type compared to all other cell types to be considered a marker gene. DESeq2-normalized counts of the BA9 expression dataset were used and cell type deconvolution was performed using the CIBERSORT R script v1.04. To test for significant differences in cell type distribution in samples from individuals with and without CUD, we performed a Bayesian estimation of the difference in means and evaluated the 95% high-density interval. The Bayesian testing was based on BEST^66^ as implemented in the R package BayesianFirstAid v.0.1. Further, an overlap analysis of DEGs in cell type markers was performed in GeneOverlap v.1.34.0^67^ using the 10-fold marker gene list from^65^ in a Fisher test.

### Functional enrichment analysis

To characterize altered biological functions related to the observed gene expression differences, we performed a gene set enrichment analysis (GSEA) for Gene Ontology (GO) terms using the gseGO function from the R package clusterProfiler v.4.6.2^68^. The DESeq2 Wald statistic defined as the log2FC divided by its standard error was used for ranking of the results. A significance threshold of q<0.05 (5% FDR) was considered statistically significant. Results of the GSEA were visualized using the emapplot function in enrichplot v.1.18.3.

### Weighted gene co-expression network analysis (WGCNA)

To identify CUD-associated co-expression patterns, we constructed co-expression modules using network analysis in WGCNA (R package v.1.72.1)^69^ and related them to CUD and other phenotypic variables available in the DBCBB cohort. Using the input matrix of normalized and variance stabilization transformed (vst) gene counts from DESeq2, a soft power threshold of 9 was estimated to achieve the criterion of scale free topology (R^2^>0.85). For the construction of networks, we used the parameters minModuleSize=10, mergeCutHeight=0.25, and maxBlockSize=36,000. The Pearson correlation of the module eigengene derived from each of the resulting n=27 co-expression modules with the phenotypes of interest including CUD was calculated to identify significant associations of the modules with phenotypes (Fig. S3A). Downstream analyses of modules significantly associated with CUD included a GO enrichment analysis using the genes assigned to the modules using the full genome as the background. Next, module genes were ranked by the product of gene significance*module membership to identify hub genes. The top 10% of module hub genes were further investigated by constructing protein-protein interaction (PPI) networks. For this, Cytoscape v.3.9.1^70^ with stringApp v.1.7.0^71^ was used. A detailed description of the PPI visualization settings in Cytoscape is found in^22^.

### Replication analysis of differential expression results

Replication analysis of CUD-associated DEGs was performed in two independent datasets where RNA-seq data from postmortem human brain tissue of the prefrontal cortex from individuals with and without CUD was available. As the first replication dataset, BA9 bulk RNA-sequencing data from N=7 individuals with CUD and N=14 control individuals originating from the National PTSD Brain Bank (NPBB)^72^ was used. Phenotypic information for the BA9 replication cohort is shown in Table S17. RNA-seq data sequenced and pre-processed as described in ^73^ was analyzed for CUD-associated differential gene expression in DESeq2 using donor age, sex, PMI, and RIN as covariates. The second replication cohort was based on a neuronal-specific RNA-sequencing dataset (GEO accession number: GSE99349) as described in ^21^. In this study, neuronal nuclei were isolated from postmortem human brain tissue of the Brodmann Area 46 subregion of the dlPFC that is laterally adjacent to BA9. Here, bulk RNA-seq data was generated from N=19 individuals with CUD and N=17 without CUD from a male mixed ancestry cohort originating from the University of Miami Brain Bank (MBB). Raw sequencing data from the replication cohort was downloaded from GEO and processed using the same analysis pipeline as in the BA9 discovery sample: 1) mapping using STAR, 2) quantification using featureCounts, and 3) DE analysis in DESeq2. For the replication analysis in MBB data, we used the same statistical model as in the discovery analysis with differential expression testing for CUD while adjusting for donor age, RIN, pH, and PMI. To explore the results, we first performed an overlap analysis of nominally significant CUD-associated DEGs (p<0.05) identified in the three datasets. Second, a targeted look-up of effect sizes (log2FC) and association p-values was performed for overlapping DEGs across datasets and for the top findings from the BA9 discovery sample, ZBTB4 and INPP5E. As an additional replication approach, we performed rank-rank hypergeometric overlap (RRHO) using the R package RRHO2 v.1.0^74^ to evaluate convergent and divergent expression patterns at the transcriptome-wide scale between studies. RRHO scores were generated based on full differential expression statistics from discovery and replication datasets followed by the evaluation of overlapping signatures between studies using the hypergeometric testing procedure as implemented in RRHO2.

### Signature-based drug repositioning analysis

With the top 150 upregulated and downregulated genes ranked by the DESeq2 test statistic from the differential expression analysis, the maximum input size in the Connectivity Map (CMap) query tool (https://clue.io/query, software version 1.1.1.43) was used to evaluate the connectivity of expression signatures (Table S8). CMap query uses the L1000 assay from the NIH LINCS project (https://lincsproject.org/) as a drug-gene expression relationship database. In L1000, expression changes for a representative set of 978 landmark transcripts are measured in response to treatment with a perturbagen such as a pharmaceutical drug^75^. In addition to the connectivity scores for individual perturbagens, CMap also provides information on perturbagen classes and a GSEA output for pathways and drug targets. Normalized connectivity scores and FDR-adjusted p-values for perturbagens and GSEA results were obtained from the CMap query tool and visualized as waterfall plots in R using ggplot2 v.3.4.2.

### Differential splicing analysis

Alternative splicing was evaluated using the annotation-free quantification approach of RNA splicing in LeafCutter v.0.2.9^28^. First, raw sequencing data were aligned to the GRCh38 reference genome using STAR with an adapted 2-pass mapping procedure. For this, the first mapping step was performed using a regular gtf-file derived genome index. The resulting splice junctions (SJ_out.tab-files) from the N=25 samples were combined and filtered so that non-canonical junctions, junctions that were supported by less or equal than 2 uniquely mapping reads, annotated junctions already covered by the gtf-file, and duplicated junctions were removed. Using the filtered splice junction output, a modified genome index was derived using STAR in genomeGenerate mode. This extended genome index containing information on gene annotation and splice junctions was used in the second mapping step resulting in the final bam-file output after mapping. Generation of junc-files, intron clustering, and differential intron excision analysis was performed as outlined by the authors of leafCutter (https://davidaknowles.github.io/leafcutter/) while including age, PMI, pH, and RIN as covariates into the Dirichlet-Multinomial generalized linear model. Default settings were used in the leafcutter_ds.R script i.e. maximum cluster size =10, minimum samples per intron = 5, minimum samples per group = 3, and a minimum coverage of 20 reads. The differential intron excision analysis results in an estimate for the change in the percent spliced in measure (ΔPSI) for each intron in a cluster and an FDR-adjusted p-value for the cluster in which the differential splicing events were detected. Differential splicing events in clusters with |dPSI| > 0.025 and an FDR-adjusted q-value<0.05 were considered statistically significant^26^. Visualizations for the differentially spliced clusters and genes were created using the leafviz extension in leafCutter. GO enrichment analysis for genes containing differentially alternatively spliced intron clusters was performed using the enrichGO function with GO “BP” ontology terms in clusterProfiler.

### Integrative gene locus analysis

Integrated visualization of functional genomics data was performed using SparK v.2.6.2^76^. Summary statistics from a meta-analysis GWAS of cocaine dependence (CD) in an EA population (N=6,378)^30^ was used to cover SNPs that are associated with CD. The EWAS summary statistics from ^22^ were used as the DNAm dataset. To prioritize the association results for visualization, SNPs and CpG sites with nominal significant association p-value (p<0.05) were filtered from the GWAS and EWAS results. ChIP-seq datasets for different activating and repressing chromatin marks were downloaded from ENCODE^77^ as deposited in the Human Reference Epigenome Matrix for dorsolateral PFC in males: ENCFF241REN (H3K4me1), ENCFF752EVS (H3K4me3), ENCFF866IWY (H3K27ac), ENCFF149DDW (H3K36me3), ENCFF784SSN (H3K9me3), ENCFF167ASN (H3K27me3). BigWig files were converted to BedGraph using the UCSC bigWigToBedGraph tool. Bam-files from the RNA-seq analysis were indexed using samtools v.1.5^78^ and then converted to BedGraph using the bamCoverage function from deeptools v.3.5.3^79^. For the genetic dataset, we performed an additional gene-based association analysis using Multi-marker Analysis of GenoMic Annotation (MAGMA)^80^. Here, we aimed to quantify the combined association of all SNPs annotated to a gene of interest with CUD as the phenotype.

### Multi-omics factor analysis

Multi-omics factor analysis (MOFA)^81^ was used to jointly analyze the DNAm and gene expression datasets in BA9 aiming for the identification of CUD-associated factors. The factor analysis framework enables an improved characterization of gene and pathway alterations across different omics datasets by investigating the contribution of each omics view such as DNAm or gene expression to a learned factor. Downstream analyses such as GSEA enable the analysis of biological functions that are associated with a factor based on the factor loading of features such as genes that contribute to the biological pathway. As the DNAm input dataset for MOFA (R package v.1.3.1), we used methylation M-values from the 20,000 most variant promoter CpG sites (TSS200 and TSS1500 annotations) under the assumption of their prominent role in regulating transcription levels of nearby genes. Methylation data was extracted for the individuals that also had expression data available (N=25). For the expression dataset, we used normalized and variance stabilization transformed counts from the 20,000 most variant genes to obtain an equal number of features in each view. The MOFA model was trained on the matched DNAm and expression data from N=25 individuals using default model options with a total of 10 factors and the training options convergence_mode = “slow”, seed = 42, and maxiter=10,000. Association of factors with phenotypes was evaluated using the correlate_factors_with_covariates function. GSEA was performed on negative and positive weights individually using the run_enrichment function based on the c5.go.bp.v2023.1.Hs.symbols.gmt gene set reference file from MSigDB^82^. Functional characterization of DNAm weights was performed by subsetting the top 2.5% of CpG sites from both sides of the weight distribution on factor 9 resulting in N=500 CpG sites with strongest positive and negative weights on factor 9, respectively. Next, GO enrichment analysis was performed in missMethyl v.1.33.1 using the full set of N=20,000 CpG sites as background.

### GO enrichment analysis of CUD-associated gene sets

Convergence of CUD association signals at the pathway level was evaluated by pathway enrichment analysis for GO terms using the enrichGO function on the GO “BP” ontology in the compareCluster functionality of clusterProfiler. A total of 10 gene lists were included in the input dataset: 1) CUD-associated CpG sites (N=394, p>0.001) from the EWAS of CUD^22^, genes in the CUD-associated WGCNA methylation modules 2) blue (N=9,201), 3) steelblue (N=390), 4) brown (N=5,268), 5) brown4 (N=205), 6) nominally significant DEGs (N=1,057,p<0.05), 7) genes in the CUD-associated WGCNA expression module yellow (N=2,517), 8) AS genes (N=98, q<0.05), 9) MOFA methylation weights factor 9 (N=983 genes based on the 2.5 and 97.5 percentiles of the weight distribution for CpG sites), and 10) MOFA methylation weights factor 9 (N=1,000 genes based on the 2.5 and 97.5 percentiles of the weight distribution for genes). Pathways remaining statistically significant after FDR correction (q < 0.05) were displayed in an enrichment map with a pie plot visualization scheme for GO terms that were repeatedly identified for the different gene lists.

## Notes

### Competing Interest Statement

The authors have declared no competing interest.

