## Supplementary Figures for "Multi-omics profiling of DNA methylation and gene expression alterations in human cocaine use disorder"

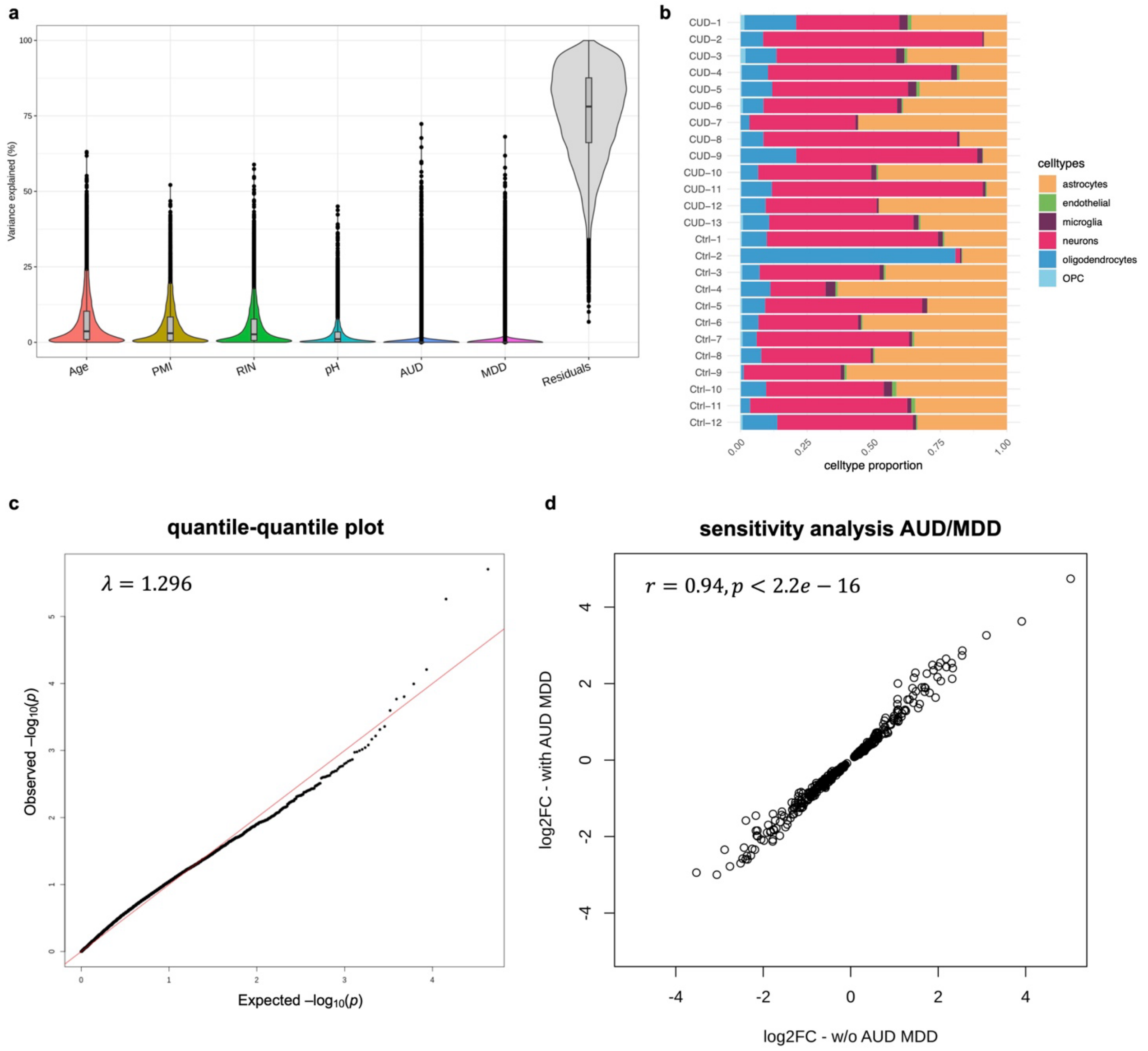


**Supplementary Figure S1 – Differential expression analysis in Brodmann Area 9**

**a** Variance partition analysis confirms the included covariates for the DE model in DESeq2 and reveals minimal explained variance for AUD and MDD. PMI=postmortem interval, RIN=RNA integrity number, pH=postmortem brain tissue pH value. **b** Cell type proportions in the N=25 postmortem human brain tissue samples from Brodmann Area 9 based on the estimation using CIBERSORT. CUD: N=13 cocaine use disorder samples, Ctrl: N=12 control samples. OPC=oligodendrocyte progenitor cell. **c** Quantile-quantile plot displaying observed vs. expected p-values in the differential expression (DE) analysis for cocaine use disorder (CUD) in Brodmann Area 9, genomic inflation factor $\lambda$=1.296. **d** Correlation of log2FC estimates for nominal significant DE genes ($p<0.05$) in the sensitivity analysis that includes alcohol use disorder (AUD) and depressive disorder (MDD) as additional covariates into the DE model. A strong and highly significant Pearson correlation between log2FC estimates was observed ($r=0.94$, $p<2.2e-16$).

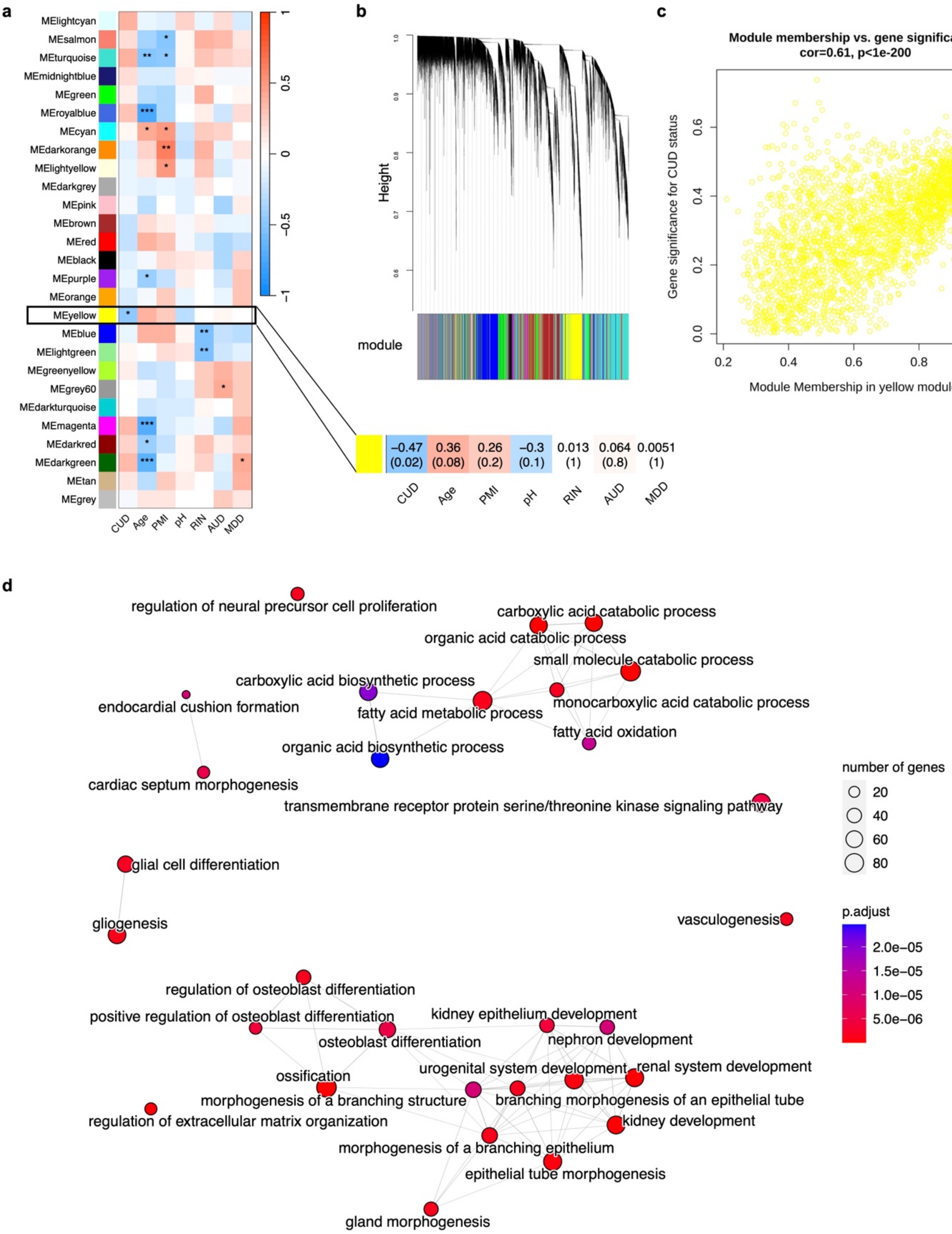


**Supplementary Figure S2 – Weighted correlation network analysis reveals a cocaine-use disorder associated co-expression module in Brodmann Area 9**

**a** Correlation of the N=27 identified co-expression modules from weighted correlation network analysis (WGCNA) with cocaine use disorder (CUD) status and other known phenotypes in the Brodmann Area 9 cohort. Pearson correlation of module eigengene with the phenotypic variable was color coded where red color indicates positive and blue color indicates negative correlation coefficients. Module yellow was significantly negatively correlated with CUD (r=-0.47, p=0.02). Significance of correlation is indicated using asterisks (*=p<0.05, **=p<0.01, ***=p<0.001). PMI=postmortem interval, pH=postmortem brain tissue pH value, RIN=RNA integrity number, AUD=alcohol use disorder status, MDD=depressive disorder status. **b** Dendrogram of genes and their assignment to co-expression modules in WGCNA. **c** Strong correlation of module membership and CUD gene significance for the N=2,517 genes in co-expression module yellow was identified (r=0.61, p<1e-200). **d** Results of the Gene Ontology (GO) enrichment analysis for the N=2,517 module yellow genes identified an overrepresentation in biological pathways related to development and fatty acid metabolism.

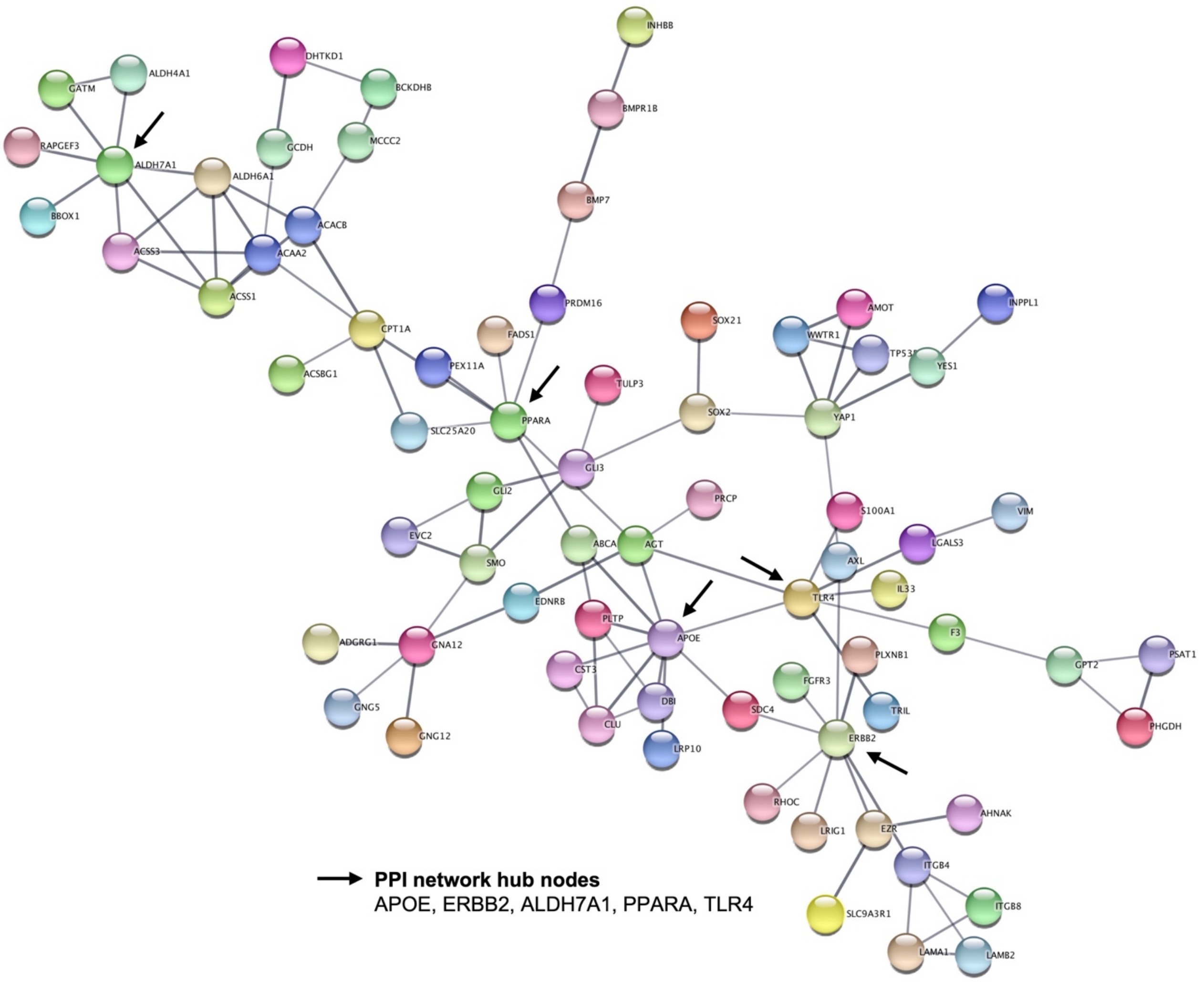


**Supplementary Figure S3 – Protein-protein interaction network of module yellow hub genes**
Protein-protein interaction STRING network of the top 10% module yellow hub genes reveals several highly connected network hub nodes such as APOE, ERBB2, and ALDH7A1, PPARA, and TLR4. Network hub nodes are highlighted by arrows.

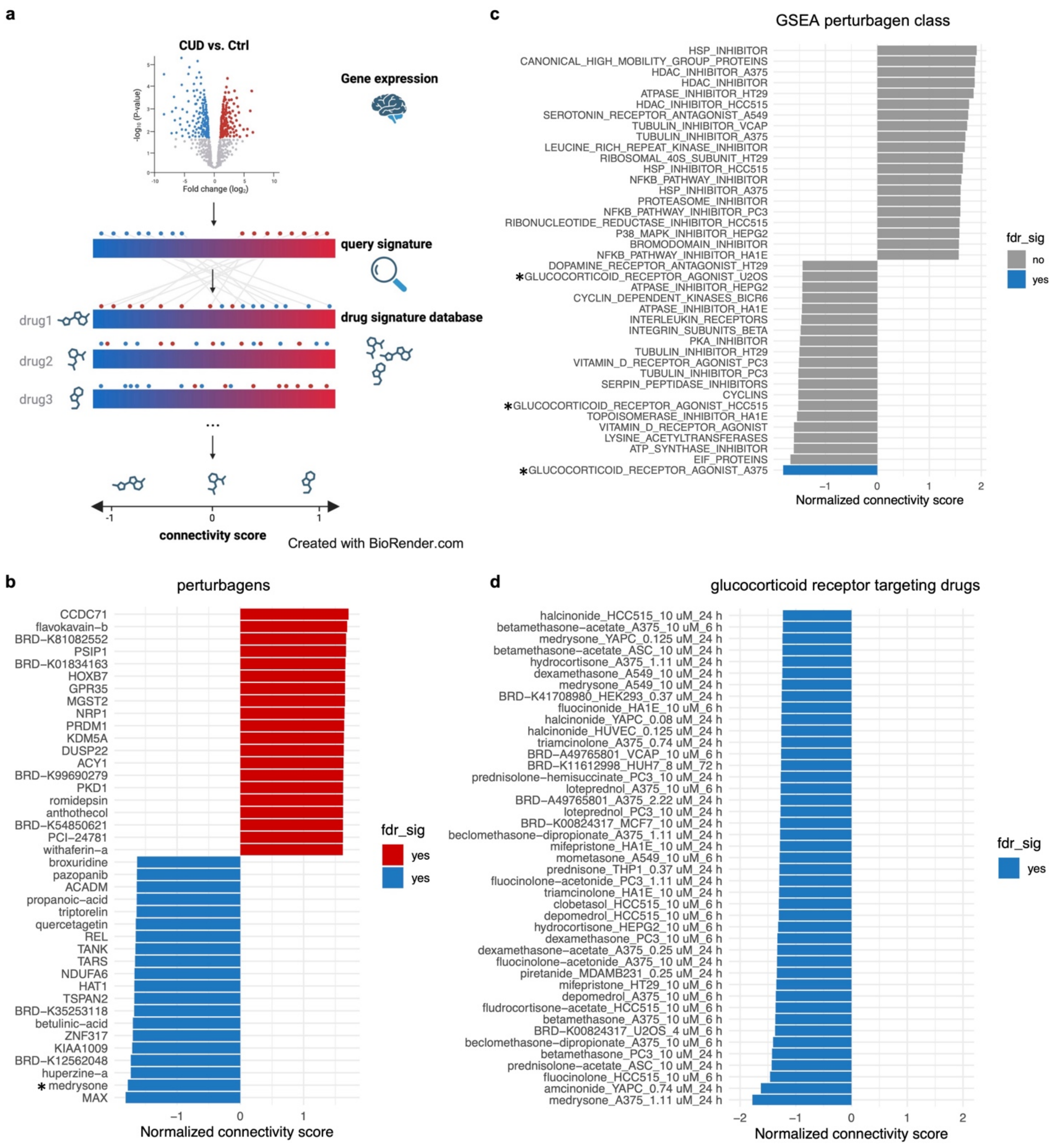


**Supplementary Figure S4 – Drug repositioning analysis based on the differential gene expression signature in cocaine use disorder suggests glucocorticoid receptor targeting drugs as a potential pharmacotherapy**
**a** Schematic overview of the drug repositioning analysis approach based on Connectivity Map (CMap) using the LINCS L1000 perturbagen reference database. The top N=150 upregulated and N=150 downregulated differentially expressed genes in cocaine use disorder were used as the query signature. **b** Results of the GSEA perturbagen class analysis from CMap. The top N=20 findings with positive and negative normalized connectivity score (NCS) are shown. Asterisks highlight findings related to glucocorticoid receptor agonists in different cell lines. **c** CMap results for individual perturbagen signatures that are most similar or dissimilar to the query signature. The top N=20 perturbagens with positive and negative NCS are shown. Asterisk highlights the glucocorticoid receptor targeting drug medrysone. **d** NCS for all glucocorticoid receptor targeting perturbagen drugs available in CMap were extracted indicating consistently significant negative NCS across cell lines compared to the query signature from Brodmann Area 9. Blue color indicates significance (fdr_sig, q<0.05) after FDR-correction for multiple testing.

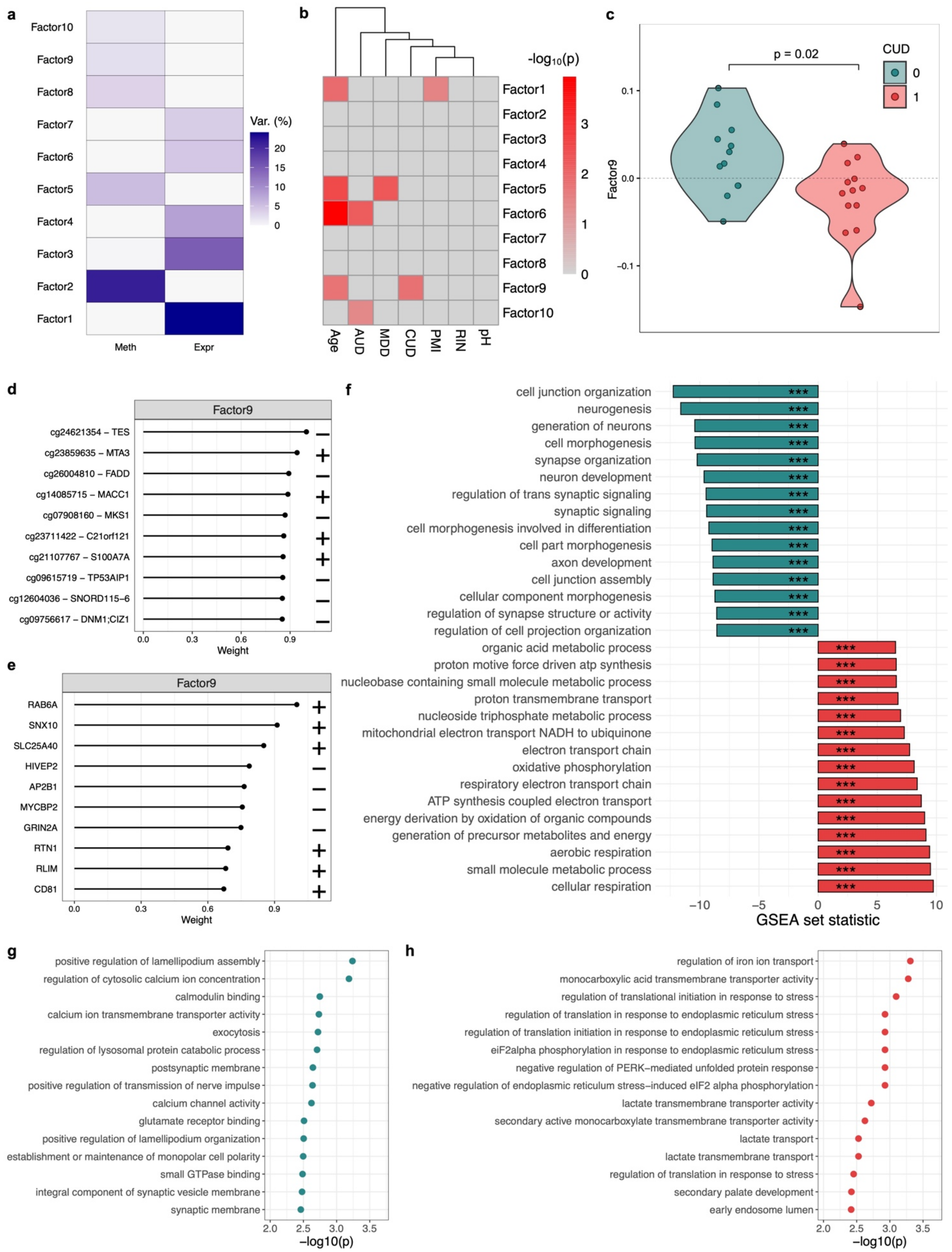


**Supplementary Figure S5 – Integrated analysis of DNA methylation and gene expression data from Brodmann Area 9 using multi-omics factor analysis provides further evidence for synaptic signaling and metabolic alterations in cocaine use disorder** **a** Variance explanation (in %) in the DNA methylation (Meth, N=20,000 most variable promoter CpG sites) and expression dataset (Expr, N=20,000 most variable genes) for each of the identified factors from multi-omics factor analysis (MOFA). **b** Association of MOFA factors with cocaine use disorder (CUD) and known covariates in the postmortem human brain cohort. AUD = alcohol use disorder, MDD/NOS = major depressive disorder/depressive disorder not otherwise specified, PMI= postmortem interval, RIN = RNA integrity number, pH = brain tissue pH value. Red color indicates statistically significant association with the phenotype ($p<0.05$). **c** Significant difference in factor 9 values was observed between CUD cases and Ctrl ($p=0.02$). Top 10 **d** CpG sites (Meth) and **e** genes (Expr) having the strongest absolute weights on factor 9. +: positive weight, -: negative weight. **f** Results of the gene set enrichment analysis (GSEA) on the expression weights on factor 9 separated by positive and negative weights. Significance estimates inside bars are FDR-adjusted q-values (***, $q<0.001$). For the DNA methylation dataset, results of the GO enrichment analysis for CpG sites with strongest **g** negative weights and **h** positive weights on factor 9 are shown.

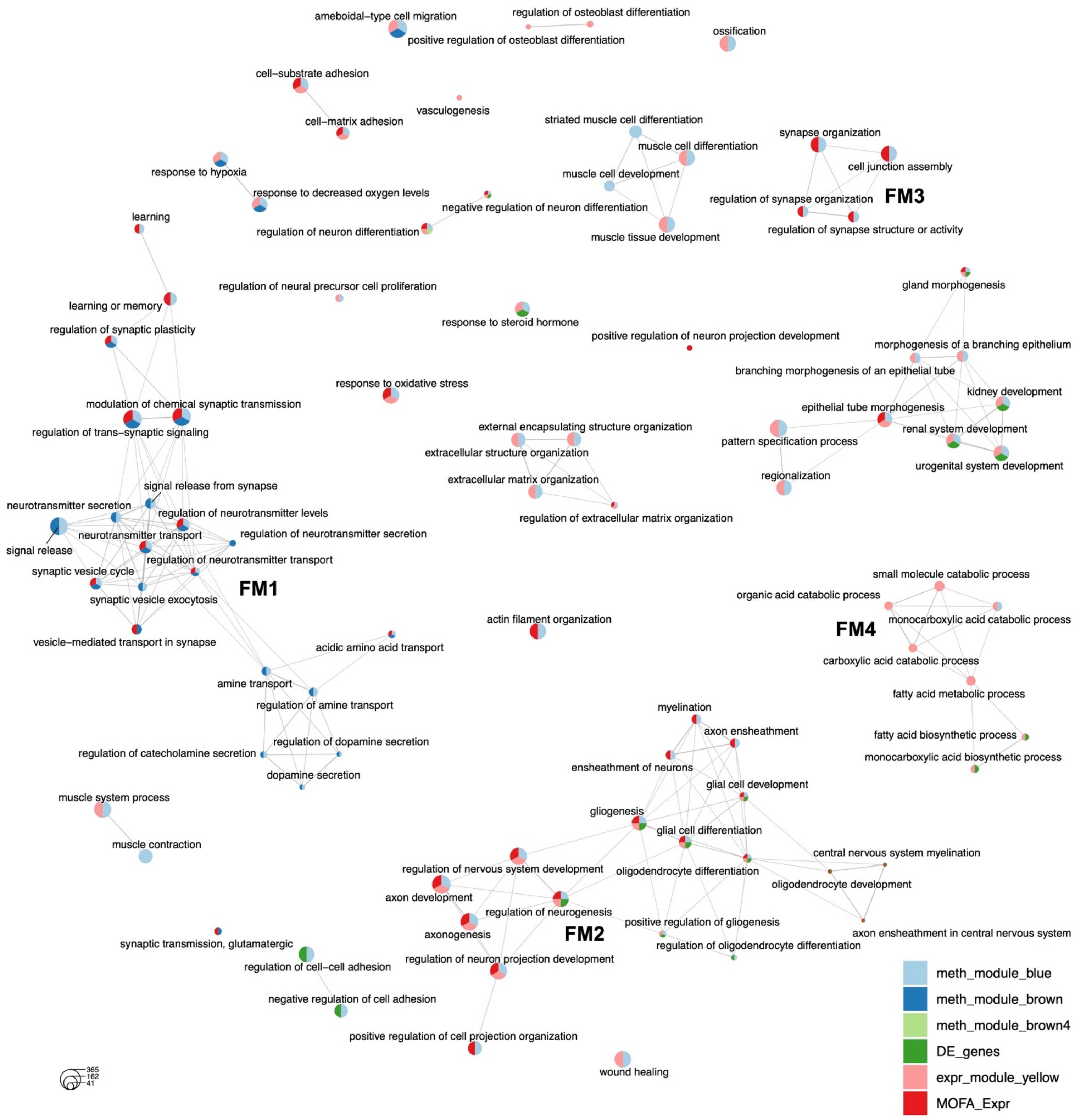


**Supplementary Figure S6 – Integrative Gene Ontology enrichment analysis based on DNA methylation and expression signatures in Brodmann Area 9 reveals multiple functional modules associated with cocaine use disorder**
Enrichment map of the Gene Ontology (GO) enrichment analysis using cocaine use disorder (CUD) associated gene lists derived from individual and integrative analyses of DNA methylation and gene expression signatures in Brodmann Area 9. Convergent evidence at the biological pathway level leads to the emergence of functional modules (FM) with interconnected GO terms where statistically significant (q<0.05) enrichment was detected for multiple gene lists.

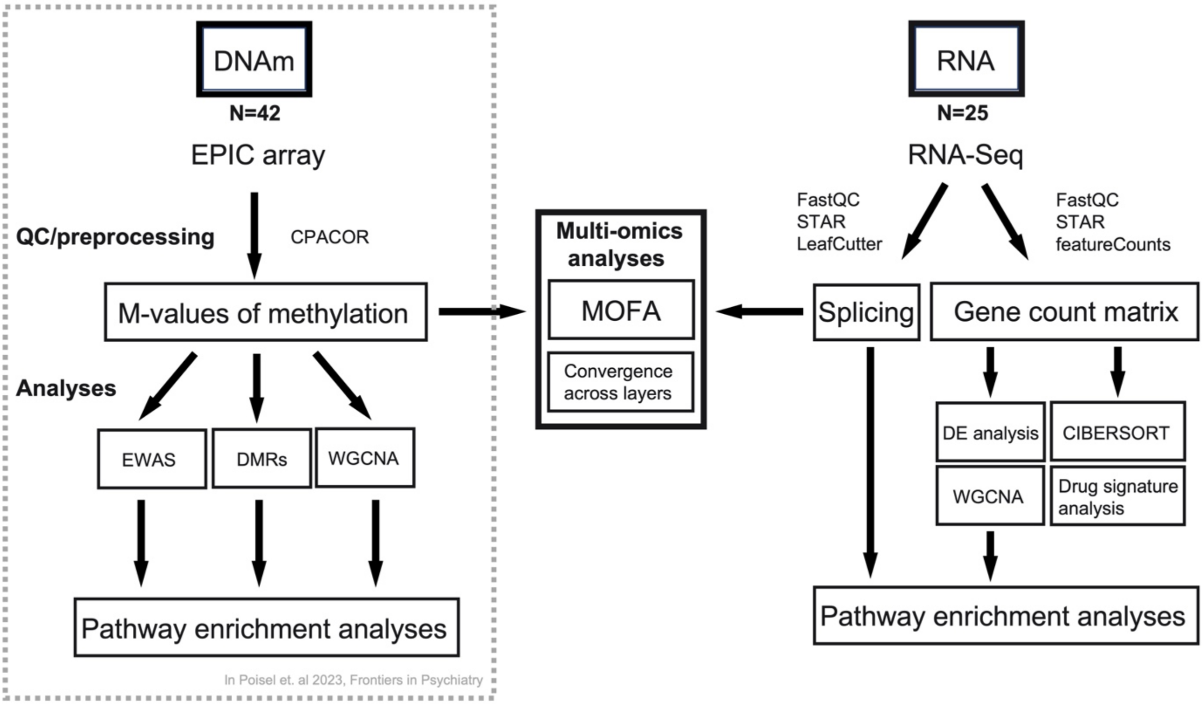


**Supplementary Figure S7 – Analysis workflow of the multi-omics analysis of DNA methylation (DNAm) and gene expression (RNA) in Brodmann Area 9 in cocaine use disorder**

Analysis of the DNA methylation (DNAm) dataset from the N=42 Brodmann Area 9 samples has been performed in Poisel et al., 2023. Transcriptome-wide gene expression analysis from RNA-sequencing in a subset of N=25 individuals of this cohort and multi-omics data integration is performed in the present study. CPACOR=Control Probe Adjustment and reduction of global CORrelation (Lehne et al., 2015), EWAS=epigenome-wide association study, DMRs=differentially methylated regions, WGCNA=weighted gene co-expression network analysis, MOFA=multi omics factor analysis, DE=differential expression. The DNAm analysis was previously published in Poisel et al., Frontiers in Psychiatry, 2023.
